# Primary Aldosteronism and Long-term Outcomes Using PAMO Definition

**DOI:** 10.1101/2025.01.15.25320635

**Authors:** Wen-Kai Chu, Yen-Hung Lin, Vin-Cent Wu

## Abstract

**Background:** Primary aldosteronism (PA) is the most common cause of secondary hypertension, associated with increased cardiovascular morbidity and mortality. This study evaluated biochemical and clinical responses to mineralocorticoid receptor antagonist (MRA) therapy and identified predictors of treatment success based on the Primary Aldosteronism Medical Outcomes (PAMO) consensus.

**Methods:** This observational study analyzed data from the TAIPAI database linked to the National Health Insurance Research Databases, including 1,305 PA patients between June 2007 and June 2021. After excluding those who underwent surgery during the study period or had relevant disease, 269 patients were classified according to their clinical and biochemical responses to MRA therapy based on the PAMO criteria.

**Results:** Among the 269 patients (mean age: 54.9±11.5 years, 40.9% male), 22.3% achieved clinical success, 55.4% partial success, and 22.3% failed. Biochemically, 56.9% of patients achieved success, 37.5% partial success, and 5.6% failed. Notably, 14.1% of patients achieved both clinical and biochemical success. After a mean follow-up period of six years, clinical success was significantly associated with a reduction in major adverse cardiovascular events (p=0.020), whereas biochemical success alone did not show the same association. Cox proportional hazards analysis demonstrated a significant association between Charlson comorbidity score (HR=1.28, p=0.003), clinical success (HR=0.29, p=0.047), and cardiovascular outcomes.

**Conclusions:** MRA therapy achieved clinical success in 22.3% and biochemical success in 56.9% of PA patients. After a mean 6 years follow-up, clinical success independently reduced MACE risk, regardless of whether biochemical success is achieved. This highlights the importance of prioritizing clinical outcomes in PA treatment.

## Background

Primary aldosteronism (PA) is a common and potentially curable cause of secondary hypertension^1,2^, characterized by autonomous aldosterone production independent of renin regulation. PA accounts for 5–10% of all hypertensive cases, with an even higher prevalence among those with resistant hypertension^2^. Patients with PA experience higher rates of cardiovascular morbidity and mortality, particularly due to hypertension and hypokalemia, compared to those with essential hypertension^3–6^. Early diagnosis and appropriate management of PA are essential for reducing long-term cardiovascular risks and improving patient outcomes.

Mineralocorticoid receptor antagonists (MRAs) are essential in the management of PA, particularly for patients who are not candidates for, or who decline, adrenalectomy^7^. However, prior research has shown that patients treated medically with MRAs face higher cardiovascular risks compared to those undergoing adrenalectomy ^5,6^. MRAs have been shown to improve blood pressure control^8^ and reduce cardiovascular events in patients with PA^9^. While MRAs effectively lower blood pressure, they do not fully offset the cardiometabolic risks of excess aldosterone, which exerts deleterious effects on the heart, kidneys, and vasculature^5,10^.

Recent research has focused on developing strategies to better predict and evaluate outcomes in PA patients treated with MRAs. One approach to assessing MRA therapy effectiveness is through the clinical response^11,12^, measured by changes in blood pressure or the defined daily dose (DDD) of antihypertensive medications, reflecting improvements in clinical blood pressure control. Another approach is biochemical response, which is typically evaluated by monitoring plasma renin activity (PRA) ^9,13^ and serum potassium levels^9,14^. PRA has been shown in recent studies to guide the management of PA and predict cardiovascular outcomes ^5,15^.

The recent release of the Primary Aldosteronism Medical Treatment Outcomes (PAMO) consensus ^16^ were developed through a rigorous Delphi consensus process involving 31 international experts in primary aldosteronism. These criteria define biochemical and clinical responses to medical treatment, providing a standardized framework for evaluating treatment efficacy. However, long-term cardiovascular outcomes using this classification remain understudied. This study aims to fill this gap by evaluating these outcomes and identifying key predictors of treatment success.

## Methods

### Patients

The TAIPAI database, which includes data from two medical centers, three affiliated hospitals, and two regional hospitals across Taiwan, initially enrolled 1,305 patients diagnosed with PA between June 30, 2007, and June 30, 2021. After excluding patients who had undergone surgery, lacked PRA or potassium data within 12 months, or had a history of myocardial infarction (MI), coronary artery disease (CAD), or atrial fibrillation prior to enrollment, 269 patients were included in the final analysis.

This study complied with the Declaration of Helsinki and received approval from the Institutional Review Board of National Taiwan University Hospital, Taipei, Taiwan (No. 200611031R). Informed consent was obtained from all participants and all procedures were conducted in accordance with the approved guidelines^17^.

### Diagnosis of PA

This study enrolled patients referred to the TAIPAI study group and underwent aldosterone-to-renin ratio (ARR) measurement for case detection of possible PA. Antihypertensive medications were discontinued for at least 21 days prior to confirmatory testing^17^, except in cases where blood pressure management required diltiazem and/or doxazosin.

Patients with an initial aldosterone-renin ratio (ARR) greater than 35 ng/dL per ng/mL/h were confirmed to have PA through saline infusion or captopril challenge tests. Further assessments, including advanced imaging or invasive procedures, were performed to subtype PA as unilateral (uPA) or non-uPA. The diagnosis of uPA was based on^17^: (1) confirmed PA, (2) imaging evidence of a unilateral adrenal adenoma or hyperplasia, (3) aldosterone hypersecretion lateralized via adrenal vein sampling (AVS) or NP-59 single-photon emission computed tomography (SPECT/CT) ^18^ if AVS was inconclusive, and (4) pathological confirmation after adrenalectomy, including CYP11B2-positive adenoma or immunohistochemical evidence of aldosterone-producing micronodules ([M]APM) ^19–21^.

### Biochemical Response Assessment

Biochemical response was evaluated by comparing baseline plasma renin activity (PRA) and potassium levels, measured on the MRA index day, with those obtained 6-12 months after initiating MRA therapy. Treatment response was classified into three categories: (1) **Success**: post-treatment potassium level ≥3.6 mmol/L and PRA >1 ng/mL/hr; (2) **Partial success**: post-treatment potassium level ≥3.6 mmol/L and PRA ≤1 ng/mL/hr; and (3) **Failure**: post-treatment potassium level <3.6 mmol/L.

### Clinical Response Assessment

Drug usage was evaluated using the Anatomical Therapeutic Chemical (ATC) classification system and the Defined Daily Dose (DDD) as defined by the World Health Organization (WHO), with DDD representing the assumed average daily dose for adults^6,22^.

According to the PASO consensus, “Prior DDD” refers to the DDD of antihypertensive medications used within the three months before MRA therapy initiation and on the MRA index day. “After DDD” denotes the DDD measured between six and nine months following MRA therapy initiation. Treatment response was classified as follows: (1) **Success**: After DDD equals zero; (2) **Partial success**: After DDD is lower than the prior DDD, or remains unchanged with a systolic blood pressure (SBP) reduction of more than 20 mmHg; and (3) **Failure**: After DDD exceeds the prior DDD, or remains unchanged without an SBP reduction of more than 20 mmHg.

### RIA and blood pressure measurement

Plasma aldosterone concentration (PAC) and plasma renin activity (PRA) were evaluated using radioimmunoassay commercial kits (ALDO-RIACT RIA kit, Cisbio Bioassays, Codolet, France) and (Ang I RIA KIt, Beckman Coulter, Immunotech, Prague, Czech Republic), respectively.

### Outcome of interests

The primary outcome of this study was the occurrence of major adverse cardiovascular events (MACE), including mortality, non-fatal myocardial infarction, non-fatal stroke, and new-onset atrial fibrillation. Demographic and baseline characteristics were collected from the TAIPAI cohort and analyzed. To ensure accuracy, patient data were cross-validated with the National Health Insurance Research Database (NHIRD) using a unique identification number, with legal authorization to track long-term outcomes^23–26^.

The Taiwan National Health Insurance (NHI) is a comprehensive program that covers ambulatory visits, hospital admissions, prescriptions, interventional procedures, and disease profiles for the vast majority of the Taiwanese population. By 2009, it provided coverage for over 99% of the population, amounting to approximately 23.12 million individuals. For each MRA user, the study tracked outcomes from the index date to the occurrence of a specified event or the end of the study, whichever came first. The follow-up period extended to December 31, 2021, ensuring a minimum of six months of follow-up for all patients.

To ensure accurate identification of outcomes, specific ICD-9-CM and ICD-10-CM codes were used to classify MACE, as these codes are highly reliable. For incident cardiac events, cross-validation was performed using records of acute myocardial infarction (AMI) ^27^, coronary artery bypass grafting (CABG) ^24^, percutaneous transluminal coronary angioplasty (PTCA), and angiography. These records were linked to procedure codes, which are integrated into the NHI reimbursement system and have been validated through auditing. Stroke diagnoses, including imaging findings and related diagnosis codes, were previously validated in this study^28,29^. New-onset atrial fibrillation diagnoses were confirmed using the corresponding ICD codes^30^.

### Statistical Analysis

To compare means among groups for continuous variables with a normal distribution, we used one-way analysis of variance (ANOVA). For categorical variables, such as sex, the presence of aldosterone-producing adenoma, diabetes mellitus, hyperlipidemia, and smoking status, we employed the Chi-square test. For non-normally distributed variables, such as hypertension duration and Charlson Comorbidity Index (CCI), the Kruskal-Wallis H test was used to compare medians. Associations between clinical and biochemical responses and clinical outcomes were assessed using Cox proportional hazards models.

A p-value of less than 0.05 was considered statistically significant, and all tests were two-tailed. Statistical analyses were performed using SPSS version 22.0 (SPSS Inc., Chicago, IL), STATA/SE 14.0 for Windows (StataCorp LP, College Station, TX), and R software version 3.4.4 (Free Software Foundation, Boston, MA).

## Results

We retrospectively identified 1,305 patients with PA. Patients were included if they received medical therapy for PA and were excluded if they had undergone adrenalectomy (n=896). Patients with cardiovascular comorbidities—such as a history of myocardial infarction (n=2), coronary artery disease (n=95), or atrial fibrillation (n=21)—were also excluded, resulting in a cohort of 291 PA patients treated with medication alone. After excluding those with missing data (n=22), a total of 269 patients were included in the final analysis (Figure 1).

**Figure 1.**
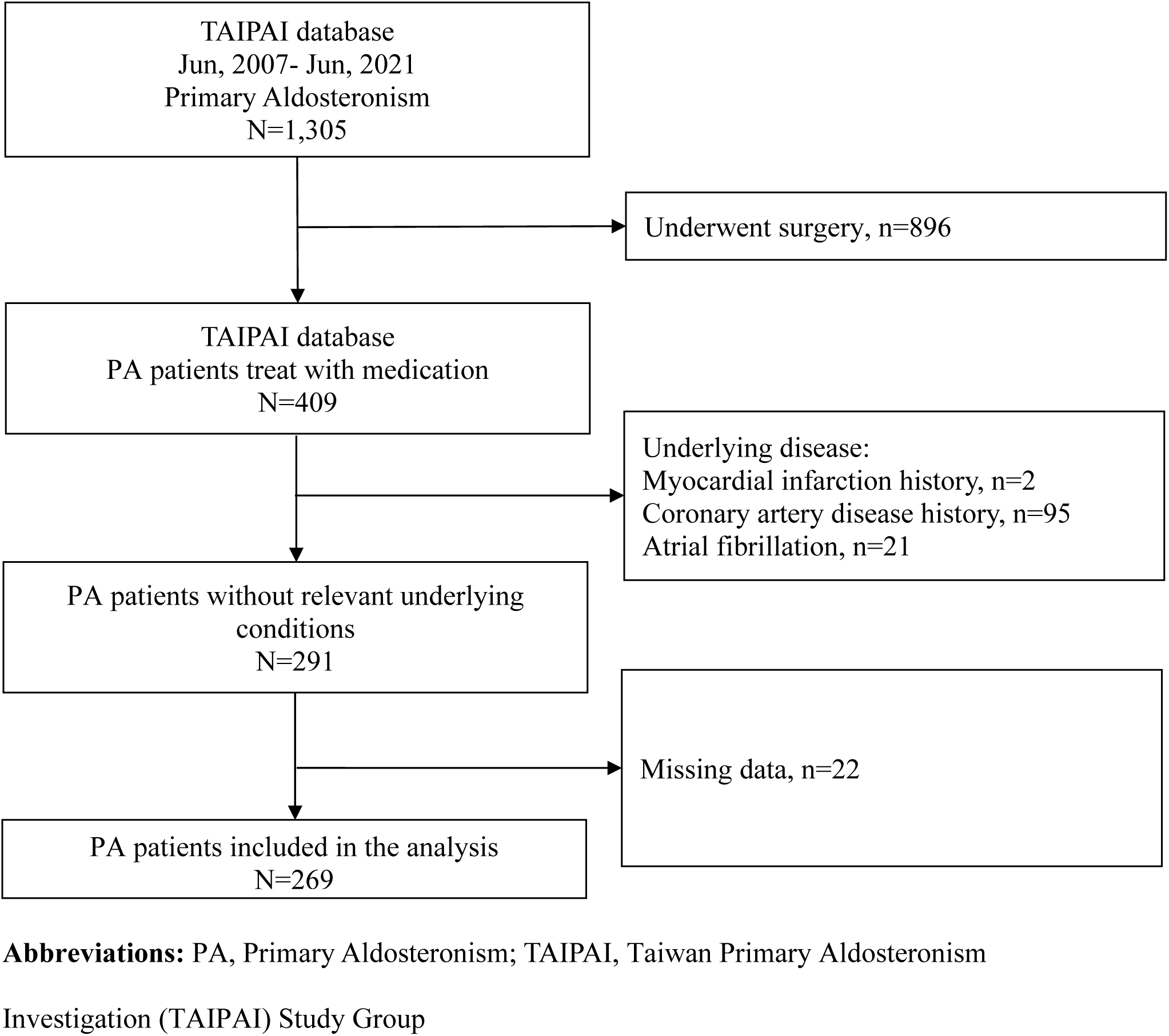
Enrollment algorithm

Within this cohort, the mean age was 54.9 ± 11.5 years, with 40.9% being male. The mean body mass index (BMI) was 25.5 ± 3.7 kg/m². Prior to treatment, the average systolic blood pressure was 151.8 ± 21.1 mmHg, and the average diastolic blood pressure was 90.4 ± 13.3 mmHg. Patients with APA constituted a significant portion of the cohort, comprising 34.6% of the total participants (Table 1).

**Table 1.**
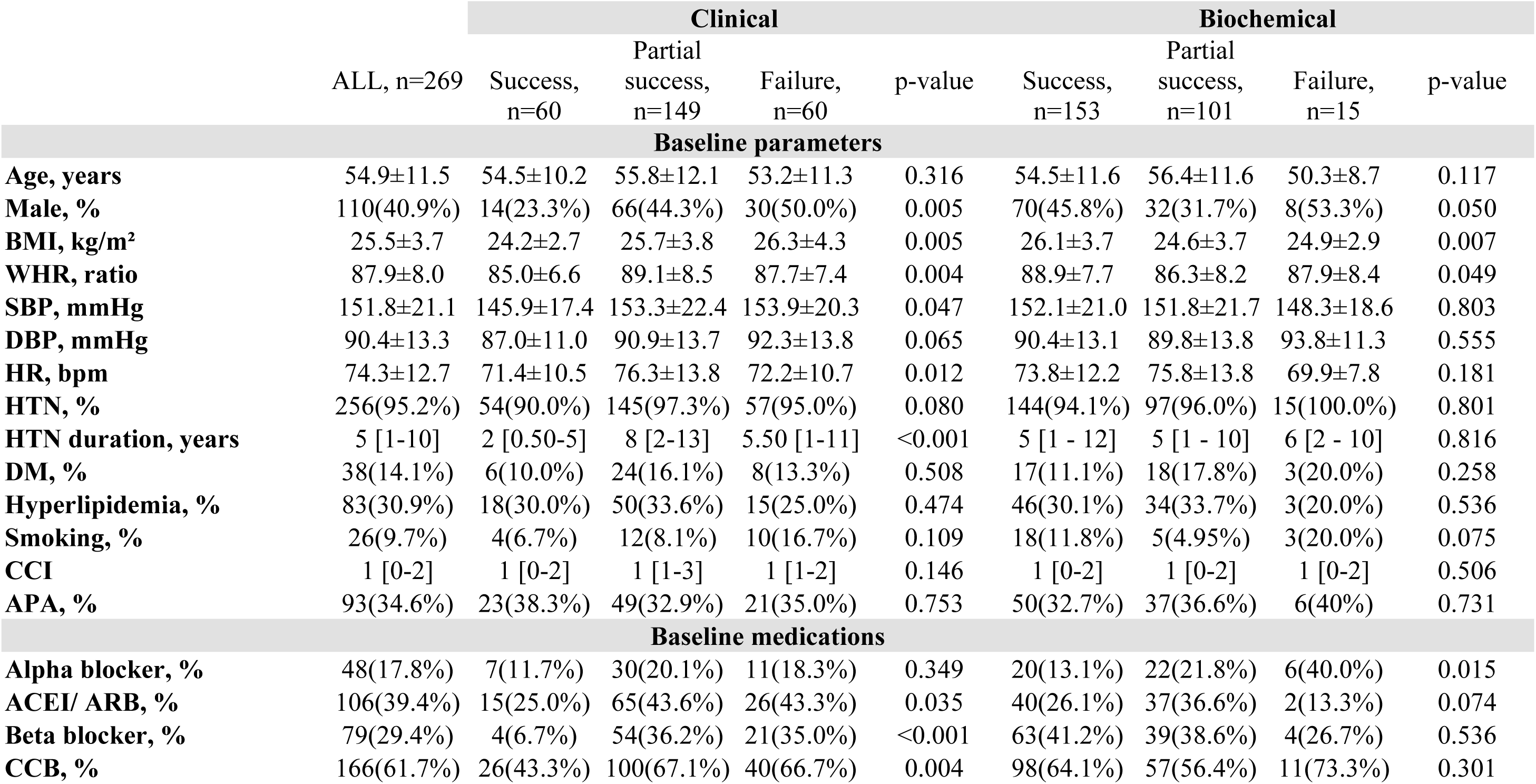
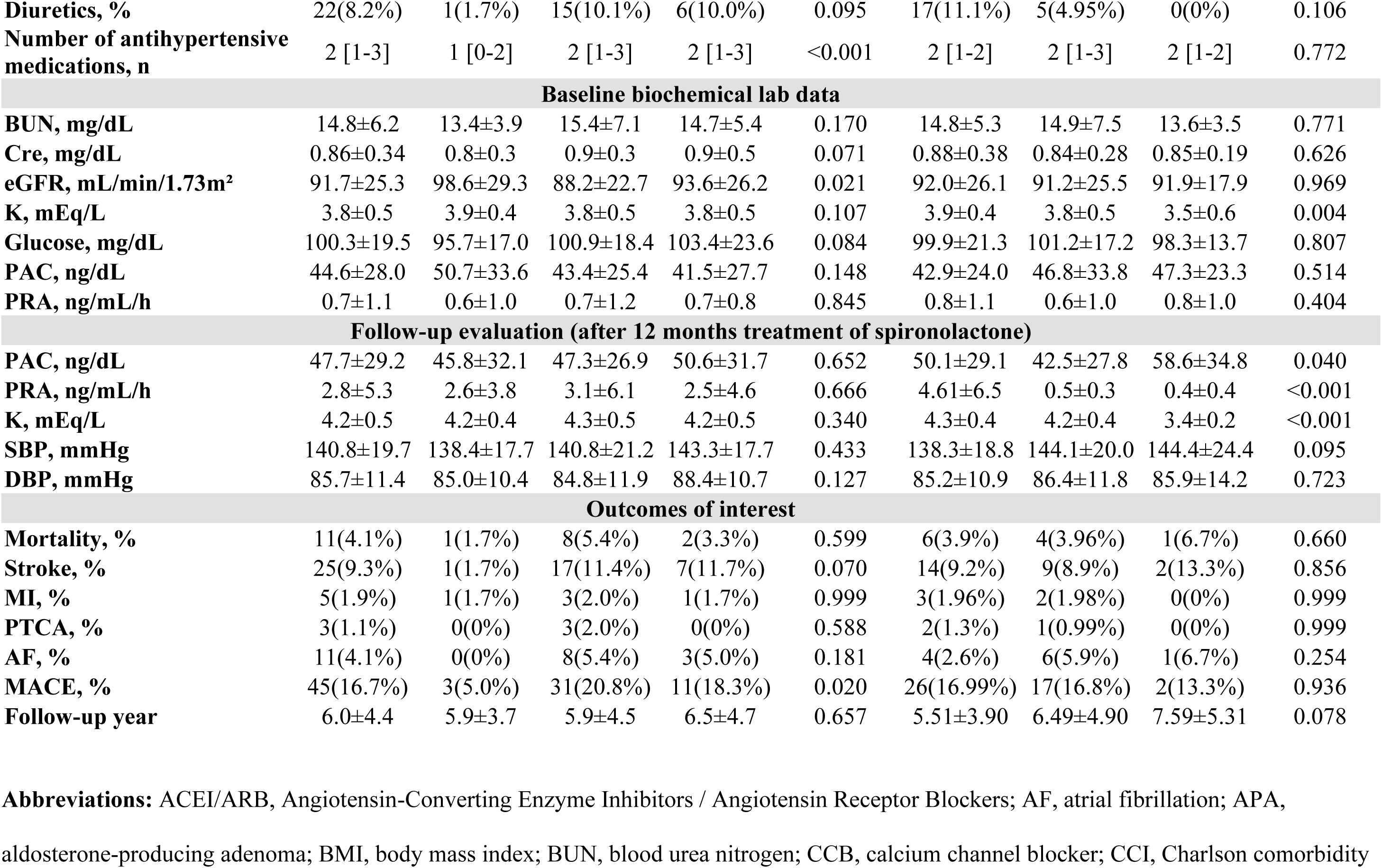
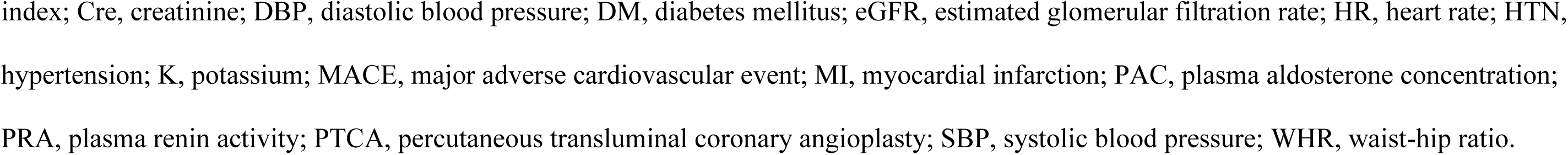
Comparison of baseline characteristics and outcomes by clinical and biochemical response in primary aldosteronism patients.

Using the consensus PAMO criteria, patients were categorized into three groups based on their clinical and biochemical responses to MRA therapy. Clinically, 22.3% (n=60) of patients were classified as having achieved success, 55.4% (n=149) as partial success, and 22.3% (n=60) as failure. Biochemically, 56.9% (n=153) of patients were categorized as having achieved success, 37.5% (n=101) as partial success, and 5.6% (n=15) as failure. In the analysis of both clinical and biochemical outcomes, 38 patients (14.1%) achieved success in both categories, while 34.9% (n=94) experienced failure in both clinical and biochemical responses (Figure 2).

**Figure 2.**
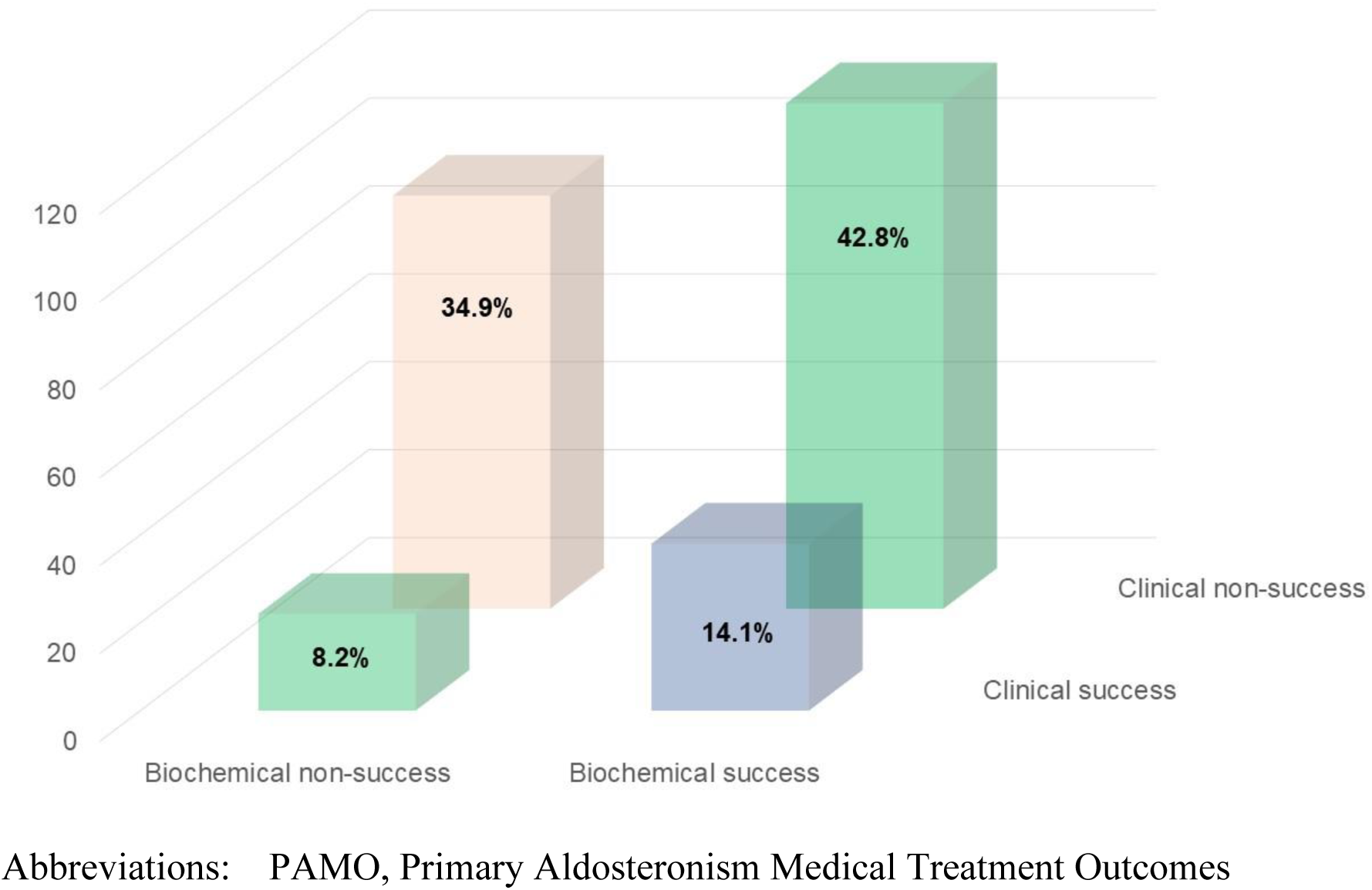
The distribution of biochemical and clinical outcomes based on the PAMO consensus

When analyzed by clinical response categories, the event rates were 5.0% (n=3) in the clinical success group, 20.8% (n=31) in the partial success group, and 18.3% (n=11) in the clinical failure group (p= 0.020). In terms of biochemical response, the event rates were 16.99% (n=26) in the biochemical success group, 16.8% (n=17) in the partial success group, and 13.3% (n=2) in the biochemical failure group (p= 0.936).

Patients who achieved clinical success were significantly less likely to be male (23.3%, p=0.005) and had a lower BMI of 24.2 ± 2.7 kg/m² (p=0.005). Furthermore, the waist-hip ratio (WHR) was lower in the clinical success group (p=0.004), and systolic blood pressure (SBP) was also significantly reduced (p=0.047).

In contrast, within the biochemical response groups, patients in the biochemical success group had a significantly higher mean BMI (p=0.007) and WHR (p=0.049). Serum potassium levels were also higher in the biochemical success group (3.9±0.4 mEq/L, p=0.004). However, there was no significant difference in SBP among the biochemical response groups (p=0.803). According to the classification criteria, PRA levels at 12 months were significantly higher in the biochemical success group (4.61±6.5 ng/mL/h) compared to the partial success (0.5±0.3 ng/mL/h) and failure groups (0.4±0.4 ng/mL/h) (p<0.001).

### Outcome of interests

During an average follow-up period of 6.0±4.4 years, the combined outcome of MACE occurred in 45 patients (16.7%) Significantly lower rates were observed in the clinical success group (5.0%) compared to the partial success (20.8%) and failure groups (18.3%) (p=0.020). However, no significant differences were observed across the biochemical response groups, with the MACE occurring in 17.0% of patients with biochemical success, 16.8% with partial success, and 13.3% with failure (p=0.936). These findings suggest that clinical response was a more significant predictor of adverse outcomes than biochemical response.

Patients who experienced the MACE were significantly older (p=0.034), had a higher prevalence of male sex (p=0.012), longer hypertension duration (p=0.005), a greater incidence of diabetes mellitus (p=0.008), and marked elevated CCI score (p < 0.001). Additionally, these patients had elevated serum creatinine levels (p=0.002) and a lower eGFR (p = 0.002). At the 12-month follow-up post-MRA treatment, systolic blood pressure remained significantly elevated in the group with adverse outcomes (p = 0.040). In contrast, other parameters, including plasma aldosterone concentration, PRA, and potassium levels, showed no significant differences between the groups (Table 2).

**Table 2.**
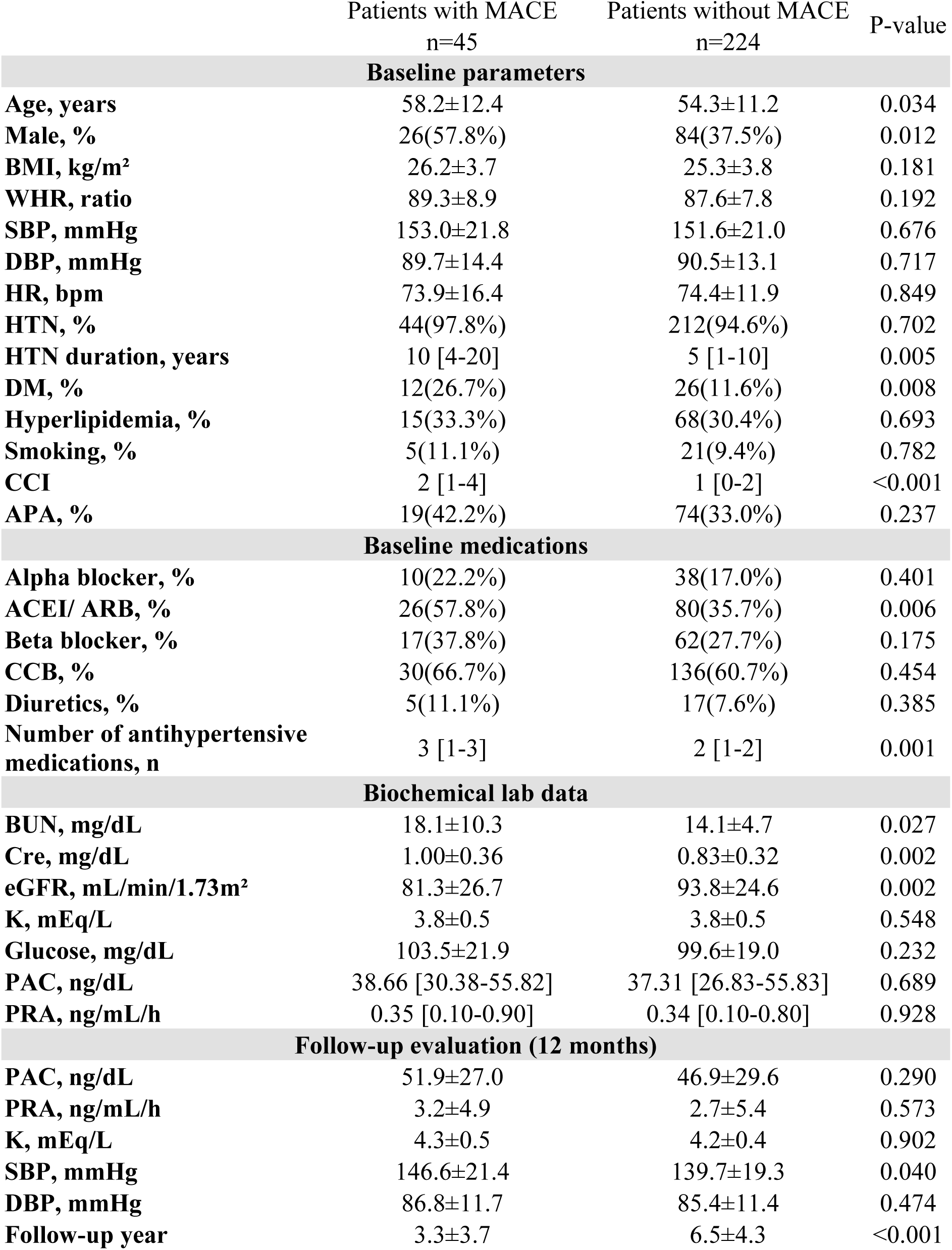
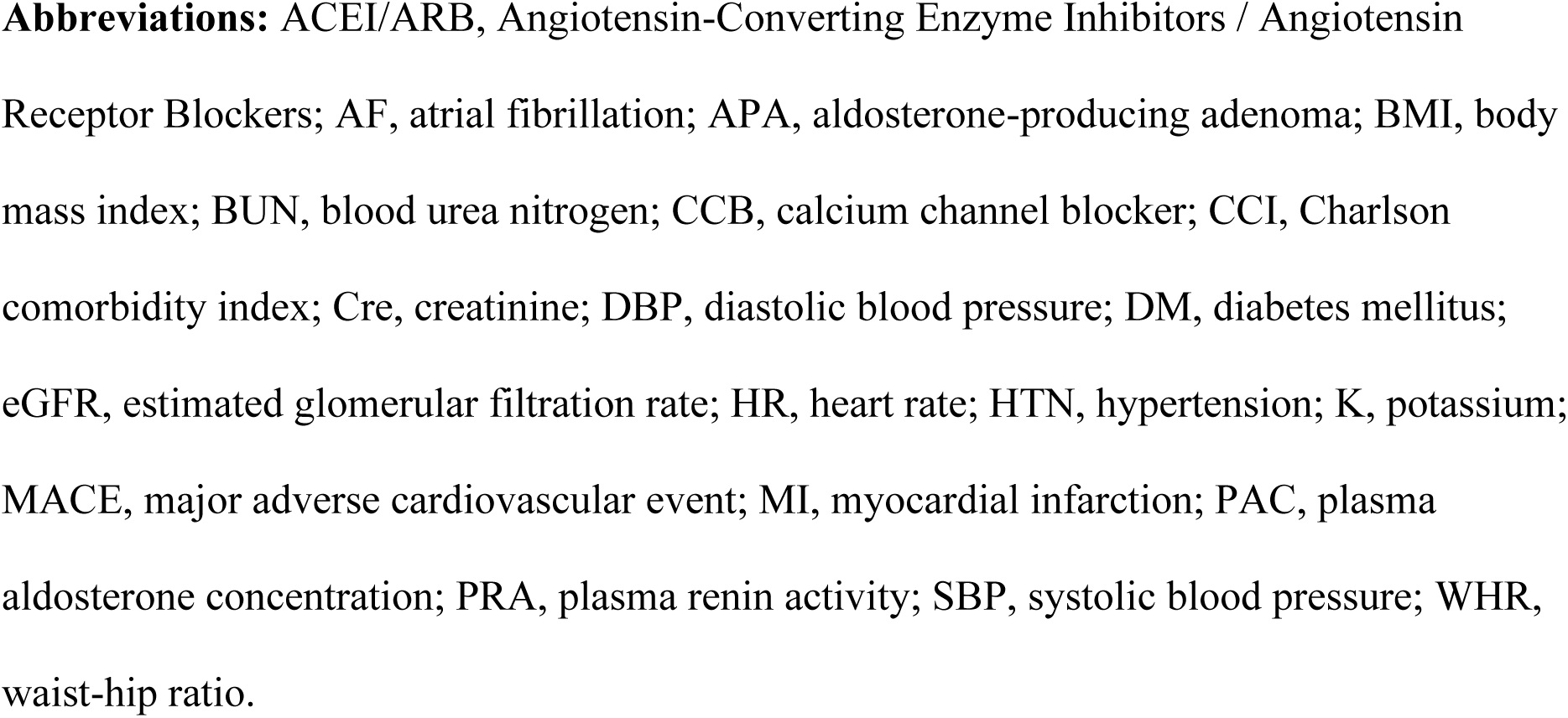
Comparison of baseline characteristics in patients with and without major adverse cardiovascular events.

The Cox proportional hazards regression analysis revealed that clinical success was significantly associated with a reduced risk of MACE (HR=0.29, 95% CI: 0.09-0.98, p=0.047). Additionally, the CCI was significantly associated with an increased risk of MACE (HR=1.28, 95% CI: 1.09-1.51, p=0.003) (Table 3).

**Table 3.** Risk factors for major adverse cardiovascular events based on Cox proportional hazard model.

| Parameter | Hazard ratio | 95% CI |  | p |
| --- | --- | --- | --- | --- |
| Age, years | 1.177 | 0.264 | 5.238 | 0.831 |
| Male | 1.622 | 0.793 | 3.318 | 0.185 |
| Body mass index | 1.335 | 0.636 | 2.806 | 0.445 |
| Systolic blood pressure, mmHg | 0.666 | 0.306 | 1.450 | 0.306 |
| Diastolic blood pressure, mmHg | 0.997 | 0.967 | 1.028 | 0.833 |
| Hypertension duration, years | 1.026 | 0.989 | 1.064 | 0.170 |
| Diabetes mellitus | 1.242 | 0.566 | 2.725 | 0.589 |
| APA | 1.242 | 0.657 | 2.347 | 0.505 |
| Creatinine, mg/dL | 1.620 | 0.756 | 3.470 | 0.215 |
| Potassium, mEq/L | 1.044 | 0.593 | 1.838 | 0.881 |
| Log PAC | 0.577 | 0.165 | 2.014 | 0.388 |
| Log PRA | 1.330 | 0.817 | 2.164 | 0.251 |
| CCI | 1.283 | 1.091 | 1.509 | 0.003 |
| Clinical success | 0.294 | 0.088 | 0.983 | 0.047 |

**Abbreviations:** APA, Aldosterone-producing adenoma; CCI, Charlson comorbidity index
; Chisq, Chi-square value; CI, confidence; Log PAC, Logarithm of plasma al dosterone
concentration; Log PRA, Logarithm of plasma renin activity

Patients were categorized into four groups based on clinical and biochemical success or non-success. In the group with both clinical and biochemical non-success, elevated serum creatinine was strongly associated with an increased risk of MACE (HR=8.69, 95% CI: 2.14-35.29, p=0.003). Additionally, the hazard ratio for non-success in both clinical and biochemical domains was 4.34 (95% CI: 1.05–18.04; p = 0.043), indicating a more than fourfold increased risk of major adverse cardiovascular events compared to patients with success in both domains (Table 4). The conditional effects plot of MACE in relation to creatinine levels demonstrated a progressive increase in MACE risk with rising creatinine levels. The plot was most pronounced in patients with clinical non-success, regardless of biochemical success or failure (Figure 3).

**Figure 3.**
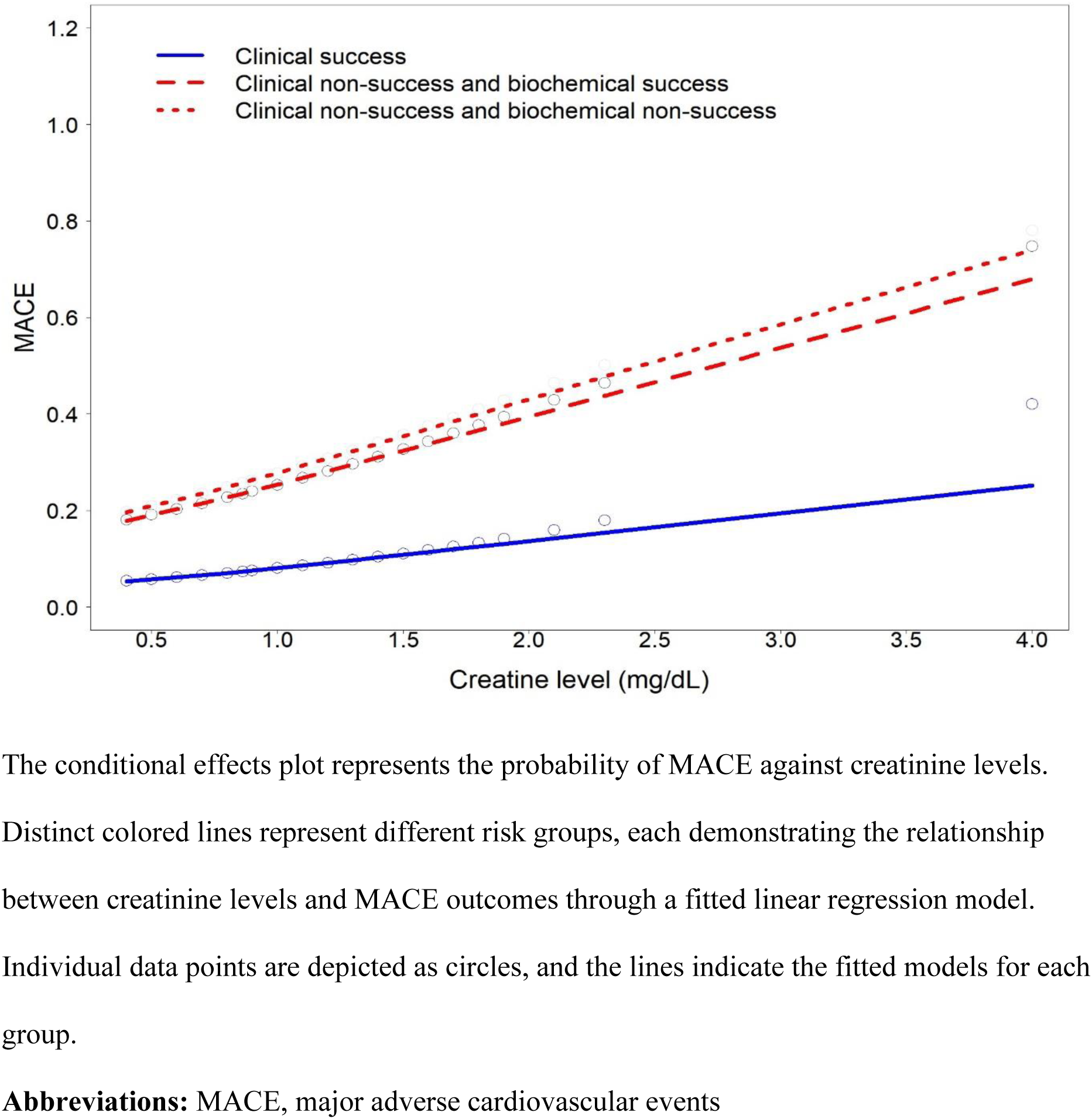
Conditional effects plot of the creatinine level on major adverse cardiovascular events

**Table 4.** Cox analysis of major adverse cardiovascular events in relation to both clinical and biochemical non-success.

| Parameter | Hazard ratio | 95% CI |  | p |
| --- | --- | --- | --- | --- |
| Age, years | 2.341 | 0.718 | 7.634 | 0.158 |
| Male | 2.307 | 0.763 | 6.978 | 0.139 |
| Systolic blood pressure, mmHg | 0.977 | 0.941 | 1.013 | 0.209 |
| Diastolic blood pressure, mmHg | 1.008 | 0.945 | 1.075 | 0.818 |
| Hypertension duration, years | 0.952 | 0.887 | 1.022 | 0.173 |
| Diabetes mellitus | 1.627 | 0.582 | 4.552 | 0.354 |
| APA | 1.350 | 0.484 | 3.768 | 0.566 |
| Creatinine, mg/dL | 8.689 | 2.139 | 35.291 | 0.003 |
| Potassium, mEq/L | 1.401 | 0.567 | 3.463 | 0.465 |
| Log PAC | 0.505 | 0.064 | 3.976 | 0.516 |
| Log PRA | 0.541 | 0.267 | 1.096 | 0.088 |
| Both clinical and biochemical non-success* | 4.343 | 1.045 | 18.042 | 0.043 |

\* compared to the reference group of patients with success in both clinical and biochemical
**Abbreviations:** APA, Aldosterone-producing adenoma; Chisq, Chi-square value; CI, confidence; Log PAC, Logarithm of plasmaaldosterone concentration; Log PRA, Logarithm of plasma renin activity

Subgroup analysis revealed no significant interaction effects with interaction p > 0.05 across various parameters, including age, diabetes status, BMI, eGFR, hypertension duration, and APA or idiopathic adrenal hyperplasia (Supplementary Table 1).

## Discussion

Our study provides a comprehensive, long-term evaluation of biochemical and clinical responses to MRAs therapy in patients with PA, utilizing the PAMO criteria. By focusing on a Taiwanese cohort with a mean follow-up of six years, this work enhances the understanding of treatment effectiveness in non-surgical PA patients.

Following MRA therapy, 22.3% of patients achieved clinical success, while 56.9% attained biochemical success. Clinical success, characterized by improved blood pressure control, was strongly associated with reduced MACE among PA patients. Importantly, an increasing creatinine level was correlated with a higher MACE risk, particularly in patients of clinical non-success, irrespective of their biochemical response.

### Risk factors associated with clinical outcomes

Clinical response, defined by improved blood pressure control and a reduction in the DDD of antihypertensive medications, may be a more significant indicator of long-term cardiovascular risk reduction^31^. Patients who achieved clinical success had a lower BMI and WHR, suggesting that reduced visceral adiposity—known to increase cardiovascular risk and impair blood pressure control—may contribute to better clinical outcomes, as supported by previous studies. ^32–34^.

Furthermore, patients with a shorter duration of hypertension may experience more favorable outcomes, as their blood vessels are less likely to have sustained significant chronic damage. In such cases, the vascular system remains more responsive to treatments, such as MRAs. Conversely, individuals with a longer history of hypertension are more prone to vascular stiffness ^35^, which increases vascular resistance and makes blood pressure management more challenging, even with effective MRA therapy ^36,37^.

### Limitation of biochemical success

In PAMO study, complete biochemical response was more frequent than complete clinical response (52.9% vs. 18.3%), consistent with our findings^16^. However, the finding that many patients achieved biochemical success without corresponding reductions in adverse outcomes suggests that current biochemical criteria may be inadequate proxies for clinical improvement. Our results indicate that while biochemical success was common, it did not consistently lead to better cardiovascular outcomes, particularly in terms of reducing MACE. In contrast, clinical success proved to be a more reliable predictor of reduced MACE risk. Patients who failed to achieve both clinical and biochemical success were more likely to experience MACE. Additionally, the conditional effects plot reinforced the importance of clinical factors, showing a significant rise in MACE risk with increasing creatinine levels, regardless of biochemical success. This highlights the need for clinical markers to better assess cardiovascular risk. Biochemical success, as defined by the normalization of PRA and potassium levels in the PAMO consensus, is based on the PASO criteria^38^.

For hypokalemia, normalizing potassium without supplementation is a primary objective that is typically achieved with MRAs^39^. Notably, the majority of contemporary PA cases do not present with baseline hypokalemia^40^. Correcting hypokalemia in patients with PA does not necessarily indicate adequate control of aldosterone levels to ensure organ protection. Indeed, even a partial reduction in aldosterone secretion may normalize serum potassium levels, raising the possibility that aldosterone activity may persist unchecked despite potassium normalization ^41^. Adjusting the dose of MRAs allows for more precise management of blood pressure, but achieving optimal aldosterone blockade can be challenging ^42^. In more severe cases of hypertension, renin-angiotensin system inhibitors (RASi) are often added to the treatment regimen. However, these medications can raise PRA, which may give the false impression of biochemical success, even if aldosterone levels are not fully controlled ^43,44^. Additionally, many PA patients and coexisting CAD are treated with beta-blockers. These beta-blockers lower renin levels, which can result in a misleading interpretation of biochemical failure, as reduced renin might be perceived as insufficient therapeutic response ^43^. Other factors, such as the timing of renin measurements and dietary sodium intake, also play a role in influencing renin levels. For example, a high sodium intake suppresses renin, while severe sodium restriction elevates it ^41,45^.

Moreover, our analysis revealed no significant differences in aldosterone or renin levels between the MACE and non-MACE groups, and neither aldosterone nor renin levels were independent predictors of MACE risk. These findings suggest that in PA patients treated with MRAs, aldosterone and renin levels may not be reliable biomarkers for predicting hypertension severity or cardiovascular risk. These confounding factors suggest that current definitions of biochemical success may obscure the true therapeutic benefit in PA patients.

In addition to achieving clinical success, patients who experienced MACE were significantly older and had a higher prevalence of male sex—both well-established cardiovascular risk factors ^46,47^. The elevated risk of MACE in male patients may be attributable to sex-specific pathophysiological differences, including heightened sensitivity to aldosterone, increased vascular reactivity, and more pronounced endothelial dysfunction, which together contribute to poorer cardiovascular outcomes. This finding aligns with previous research showing that a significantly higher percentage of women achieved complete clinical response compared to men following targeted treatment (26.4% vs. 10.6%, p<0.001) ^16^. Both chronic hypertension and diabetes promote endothelial dysfunction and vascular remodeling, resulting in increased arterial stiffness and resistance, which complicates blood pressure management, even with MRA treatment. ^48,49^.

In our study, there was no significant difference in MACE risk between patients with APA and those with idiopathic adrenal hyperplasia, despite prior evidence linking APA with a more severe clinical profile and higher aldosterone secretion. This finding may be attributed to the surgical management typically applied to APA patients less comorbidities, leaving a subset of APA patients with severe comorbidities disease managed non-surgically.

### Study Strength and Limitation

In the absence of large, robust clinical trials assessing hard outcomes, treatment strategies for PAMO have primarily been shaped by physiological principles and expert consensus^44^. This reliance on expert opinion highlights the need for more concrete data to inform treatment decisions. Our study addresses this gap by utilizing comprehensive health insurance registry data, with minimal missing information. This use of real-world evidence provides valuable insights that can help refine treatment strategies and improve patient outcomes, especially in conditions like PAMO, where clinical trials are scarce. We emphasizes the importance of real-world data in supplementing traditional clinical research, particularly when high-quality, large-scale trials are lacking.

Several limitations of this study should be acknowledged. First, the retrospective design inherently limits causal inference and may introduce selection bias. Additionally, the average follow-up period was relatively short, which may have been insufficient to capture long-term cardiovascular outcomes. The relatively small sample size may have limited the study’s power to detect significant differences in certain subgroups or outcomes. This limitation arises from our policy of encouraging patients to undergo adrenalectomy, given its superior clinical outcomes compared to medical treatment^50^. It is also important to note that data were obtained from health insurance registry records, where diagnostic procedures were not as tightly controlled as they would be in structured, single- or multi-center clinical studies. Nevertheless, the use of real-world data, including comprehensive medication records, enabled us to analyze long-term outcomes in a large population of patients with primary aldosteronism, utilizing ICD diagnostic codes and CPT coding within a cohort of 23 million individuals. We believe this big-data approach provides valuable insights into the understanding of PA.

Moreover, the study population was limited to Taiwanese individuals, which may restrict the generalizability of our findings to other ethnic or racial groups. Genetic and environmental factors may influence treatment response and disease progression, necessitating caution in extrapolating these results to other populations. Additionally, we were unable to distinguish between patients with bilateral adenomas and those with unilateral hyperplasia, as these conditions are often challenging to differentiate due to the presence of micronodules. Despite the low incidence of these subgroups, our findings remained robust across various analytical models, supporting the strength of our conclusions. The diagnostic reliability of PA and its comorbidities was thoroughly validated within our cohort, enhancing the credibility of the results. However, further studies with more precise subgroup identification and longer follow-up periods will be necessary to confirm these findings in broader populations.

In conclusion, using the PAMO criteria, our study found that 22.3% of PA patients achieved clinical success, and 56.9% achieved biochemical success. Importantly, clinical response emerged as a more robust predictor of long-term cardiovascular outcomes after a six-year average follow-up. These results underscore the need for a more comprehensive evaluation of treatment efficacy in PA patients, emphasizing that clinical responses should be considered. This nuanced approach is essential for improving cardiovascular outcomes and guiding more personalized treatment strategies in PA with MRA management.

## Data Availability

The datasets generated and analyzed during the present study are available from the corresponding authors on request.

## Declarations

### Ethics approval and consent to participate

This study complied with the Declaration of Helsinki and received approval from the Institutional Review Board of National Taiwan University Hospital, Taipei, Taiwan (No. 200611031R). All experimental protocols research was approved by the Institute Research Ethical Committee of National Taiwan University Hospital (NTUH) (http://doi.org/10.6084/m9.figshare.21730985).

All methods were carried out in accordance with relevant guidelines and regulations. Informed consent was obtained from all participants or their legal guardians.

### Consent for publication

Not Applicable

### Authors’ contributions

Wen-Kai Chu: Formal analysis, Visualization, Writing - original draft

Yen-Hung Lin: Data curation, Formal analysis, Writing - review & editing.

Vin-Cent Wu: Conceptualization, Data curation, Formal analysis, Methodology, Resources, Supervision, Validation, Visualization, Writing - review & editing.

## Acknowledgements

We would like to thank the National Taiwan University Hospital, Taiwan’s National Health Research Institutes, Taiwan’s Ministry of Science and Technology. We thank Membership of the Taiwan Primary Aldosteronism Investigation (TAIPAI) Study Group (http://doi.org/10.6084/m9.figshare.21669929).

